# CoViD-19 rRT-PCR Testing Capacity in Ghana; Indications of Preparedness for Marburg virus Outbreak?

**DOI:** 10.1101/2022.10.11.22280953

**Authors:** Eric NY Nyarko, Justice Kumi, Emmanuel K Ofori, Richmond O. Ateko, Michael Appiah, Afua B. Adjei, Derrick N.D. Dodoo, Broderick Yeboah Amoah, Sandra ANA Crabbe, Ebenezer K Amakye, Seth D Amanquah, Christian Obirikorang, Henry Asare-Anane

## Abstract

**Introduction:** Ghana, as of July 2022, has had 168,350 Real-Time Reverse Transcription-Polymerase Chain Reaction (rRT-PCR)-confirmed cases of CoViD-19 infections and 1,458 deaths. Besides, 2 cases of Marburg virus diseases (MVD) were confirmed in the country within the same month. Both CoViD-19 and MVD require rRT-PCR for diagnosis, however, rRT-PCR facilities are scarce in Ghana – especially, hitherto, the CoViD-19 pandemic. The objectives of this study were to assess the current testing capacity of CoViD-19 rRT-PCR in Ghana, and to make some recommendations in case of an MVD outbreak, or recurrence of the CoViD-19 pandemic.

**Methods:** The study was cross-sectional. Questionnaires were administered to 100 health professionals actively involved in the testing cycle of CoViD-19 across rRT-PCR testing institutions. Responses with regards to CoViD-19 rRT-PCR testing, biosafety, and relationship with Surveillance Outbreak Response Management and Analysis System (SORMAS), PanaBios and Zipline, were obtained for 2020-through-2022. The responses were analyzed with Microsoft Excel office-365 and SPSS v.23.

**Results:** Thirty-five (35) of the 53 testing institutions were in the Greater Accra Region, but none in seven (7) regions of the country. Many (49%) were privately owned. Nine (9) different professionals were involved in rRT-PCR testing. The testing institutions increased from 2 (in March 2020) to 53 by June-ending 2022, and most (90%) had Biosafety Cabinet class II (BSCII). PPEs were inadequate between march and June, 2020 (25%), but enough (100%) by June 2022. Zipline, SORMAS, and PanaBios, respectively, saw transactions from 28%, 81%, and 77% of the institutions.

**Conclusion:** Ghana is adequately resourced for recurrence of CoViD-19, or any MVD outbreak, in terms of diagnosis with rRT-PCR. However, the country needs redistribution of these testing resources, expand the services of Zipline and SORMAS, satisfy additional biosafety requirements for MVD testing and equip over 180 GeneXpert facilities to help in accessible and affordable testing.

## Introduction

The CoViD-19 pandemic has infected more than 574 million of the world’s human population, with a death rate of 6.3 million (1.1 %) as of 31^st^ July, 2022.(1) In Ghana, there has been 168,350 Real-Time Reverse Transcription-Polymerase Chain Reaction (rRT-PCR) confirmed cases with 1,458 deaths (0.86%).(2) At the peak of the pandemic, especially within the 2nd and 3rd quarters of 2020, the importance of rRT-PCR testing to clinical management, contact tracing, isolation of infected persons and quarantine of same, were extremely important. Countries such as Germany and North Korea, were able to manage the CoViD-19 pandemic effectively, because they were able to identify and organize their testing capacity earlier.(3) In Ghana, there was a scarcity of nucleic acid amplification tests (NAAT) infrastructure, particularly rRT-PCR equipment, and biosafety cabinets (BSC) class II, capable of containing class 2 and 3 dangerous viruses, making testing within appropriate turn-around times (TAT) nearly impossible

Currently, Ghana is within the peak and post-pandemic phases, and the demand for PCR testing may continuously decline, despite the possible widespread availability of CoViD-19 PCR testing (i.e. September 2022). Moreover, on the 1^st^ of July, 2022, two (2) cases of Marburg virus (MV) infections resulting in Marburg virus diseases (MVD), were confirmed in the Ashanti region in Ghana.(4) Marburg virus diseases is a highly infectious disease that causes haemorrhagic fever.(5) The gold standard for diagnosis require rRT-PCR as the SARS-CoV-2 virus which causes the CoViD-19 infectious disease. However, blood sample is required and occasionally buccal, nasopharyngeal or oropharyngeal samples - required for CoViD-19.(5) Though they have similar early stage clinical presentations,(6), MVD is haemorrhagic, has a high case fatality rate(CFR) averaging 50% (compared with <5.0% for CoViD-19). Marburg virus diseases has no specific antiviral agent, unlike CoViD-19, it has no vaccine, and requires Biosafety cabinet class III (BSCIII) during testing.(5–7) This study was done to assess the CoViD-19 pandemic’s testing response infrastructure - including rRT-PCR, Sample transport, Health and Laboratory Information/Management Systems (HLIMS); and Biosafety equipment - build-up (or capacity) so far in Ghana, in case of post-peak CoViD-19 recurrent or MVD outbreak. The most popular HLIMS employed in this pandemic was the SORMAS and transportation of samples outside major suburbs, towns and villages were mostly done by a drone service company called Zipline. Zipline played a significant role in the management of the pandemic – especially when the pandemic was at its peak. These include transport of samples between sample collection and testing sites, and delivery of pharmaceutical and other medical supplies from its 6 main stations. These stations were the Omenako, Mpanya and Anum in the Eastern Region, Vobsi in the North East region, Sefwi Wiawso in Western North and Kete Krachi in the Oti region.

## Methods

The study design was a cross-sectional survey conducted across the 16 regions of Ghana between May and July 2022. Questionnaire was initially prepared and reviewed by 10 different professionals who were actively involved in the entire testing cycle, during the pandemic. The final structured questionnaire was administered to 100 health professionals who are actively involved in different aspects of the CoViD-19 pandemic (especially in testing) across 53 CoViD-19 rRT-PCR registered institutions, consisting of 58 centres (ie 3 of the institutions had 2 or 3 testing centers) in Ghana. Seventy-two (72) of the health professionals who were able to respond to at least 21 (75%) of the 28 questions relevant to the objectives of the study were included in the data analysis. The response rate (or ‘better’ adequate response rate) was 72%. The respondents included Medical Laboratory Scientists (MLS), Research Scientists, Laboratory Technologists and Technicians, Microbiologists, Fulfilment Operations Professionals (FOP), Laboratory Physicians, Lecturers, Molecular Biologists and Veterinary Officers. Others included a Data Entry Clerk, and a PCR Technician. Responses to questions such as the location of the testing facility, rRT-PCR equipment in-use, date of commencement of rRT-PCR testing, biosafety, costs of reagents and tests, turn-around-times (TAT) of tests, the average number of tests per day, transactions with Surveillance Outbreak Response Management and Analysis System (SORMAS), PanaBios and Zipline were obtained for 2020, 2021 and 2022. The responses were entered into Microsoft Excel office-365 and analyzed, together with SPSS v.23.(8,9)

### Patient and Public Involvement (PPI)

This study did not include patients, neither did it collect health or medical data from the participants.

## Results

Thirty-five (66%) of the 53 rRT-PCR testing institutions were found in the Greater Accra Region, 5(9%) in the Ashanti, 3 (5%) in western, 2(4%) in each of Eastern, Northern, Savannah and Upper East; while Oti and Volta had 1(2%) each (***Figure 1a***). None was found in Ahafo, Brong Ahafo, Bono East, Central, North-East, Upper West, and Western North regions of the country. Many, 26 (49%) of the institutions are privately owned, 21 (40%) are publicly owned, 5 (9%) are public-private-partnerships (PPP) and 1 (2%) is a non-governmental organization (an NGO) owned (***Figure 2***).

**Figure 1a:**
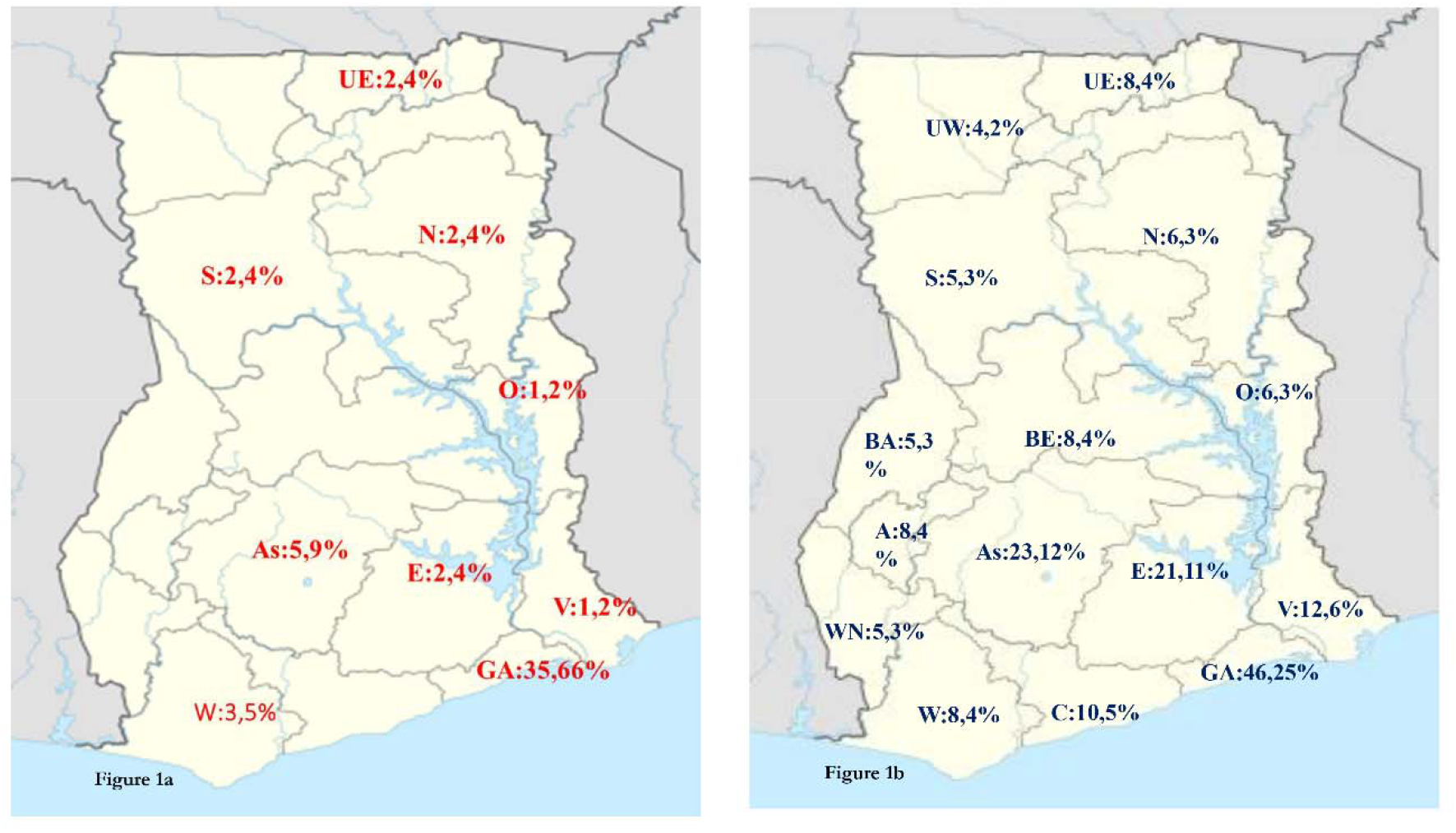
Distribution of rRT-PCR in Ghana without most of the GeneXpert Infrastructure, and Figure 1b : Distribution of rRT-PCR in Ghana upon addition of the GeneXpert Infrastructure A: Ahafo, As: Ashanti, BA: Brong Ahafo, BE: Bono East, C:Central, E:Eastern,GA:Greater Accra, N:Northern, O:Oti, S:Savannah,UE:Upper East, UW: Upper West, V:Volta, W: Western, and WN: Western north regions. NB: 17, 9% are distributed either in Teaching Hospitals, Research Institutions or Islands

**Figure 2:**
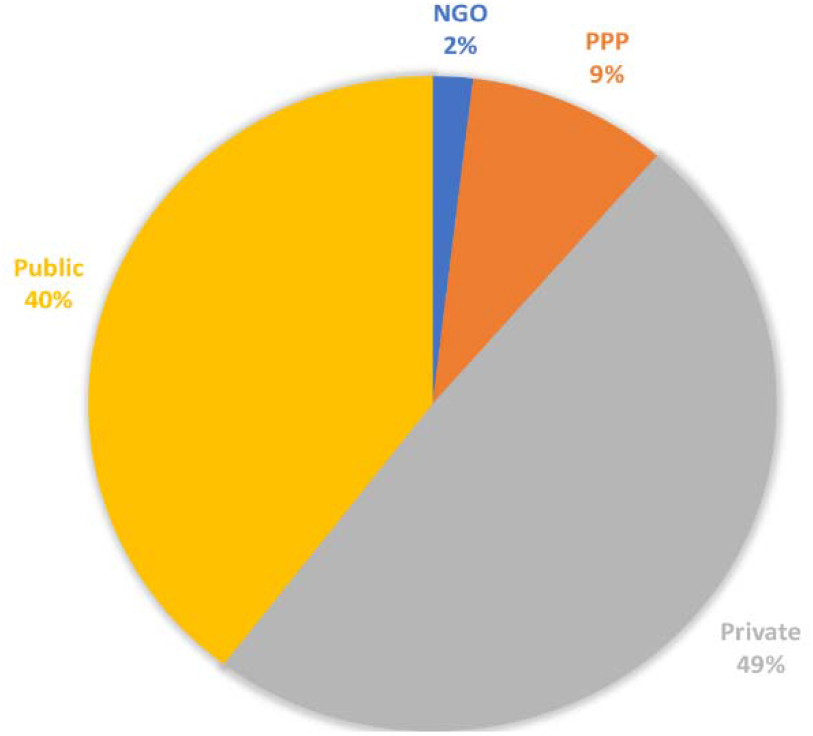
Type of ownership of rRT-PCR testing institutions. NGO: Non-Governmental Organization; PPP: Private public partnership.

Nine (9) different professionals and experts were involved in rRT-PCR testing (i.e. bench work); including Medical Laboratory Scientists (MLS), 30 (59%), Research Scientists, 7 (13%), 3 (6%) each of Laboratory Technicians and Microbiologists, 2 (4%) each of Laboratory Physicians, Molecular Biologist and Veterinary officers, and 1(2%) each of PCR technician and Laboratory technician (***Figure 3***). Other professionals who were involved (indirectly) in the testing cycle (and were included in the study) were 2 fulfilment operation professionals (FOP) from Zipline and a data entry clerk from SORMAS.

**Figure 3:**
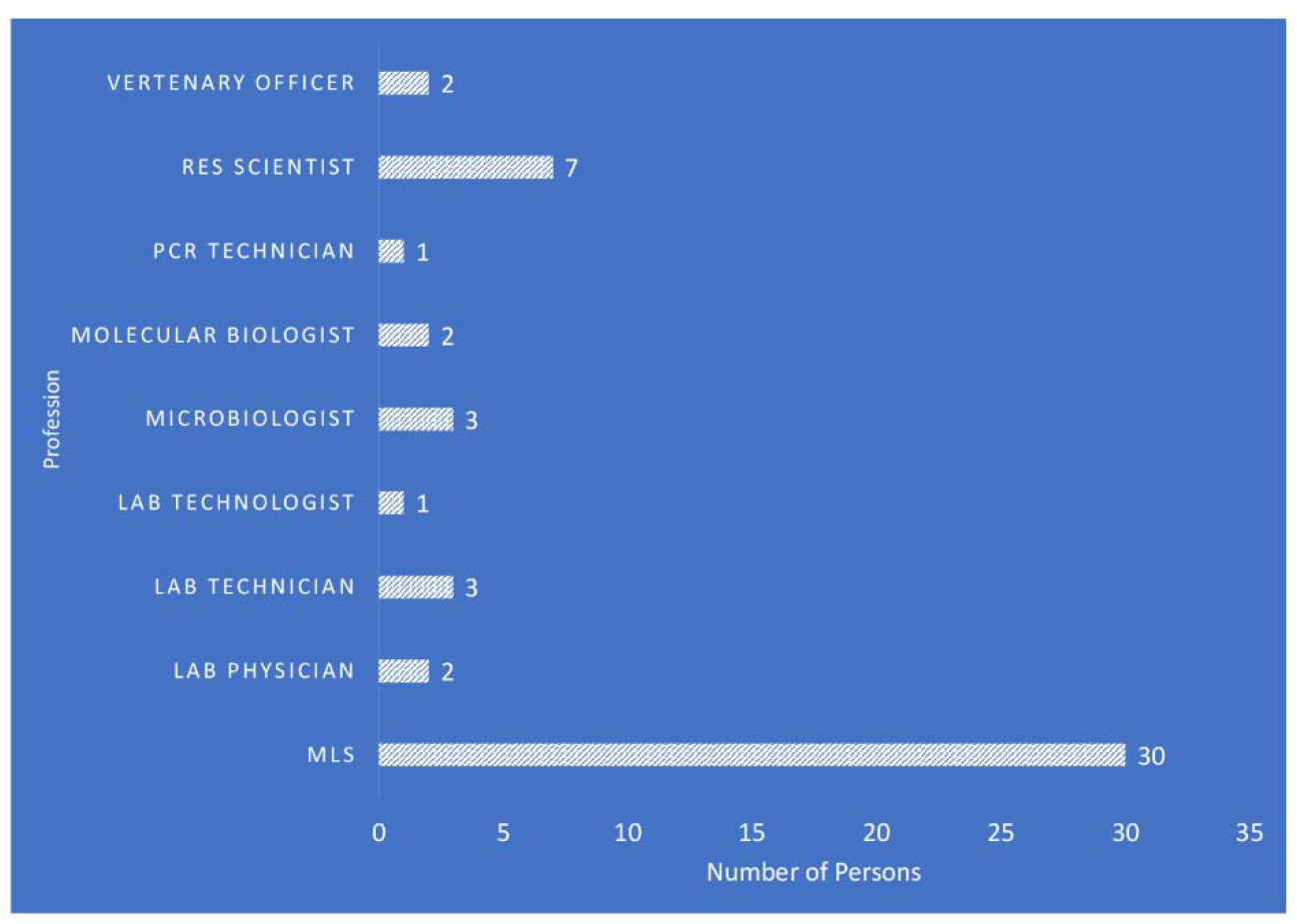
Categories of Professional expertise involved in rRT-PCR direct testing (bench work)

The number of rRT-PCR testing institutions increased from 2 in march 2020 to 35 by October 2020, 40 by February, 2021; 48 by June 2021; 51 by October 2021 and 53 by June 2022 (***Figure 4***).

**Figure 4:**
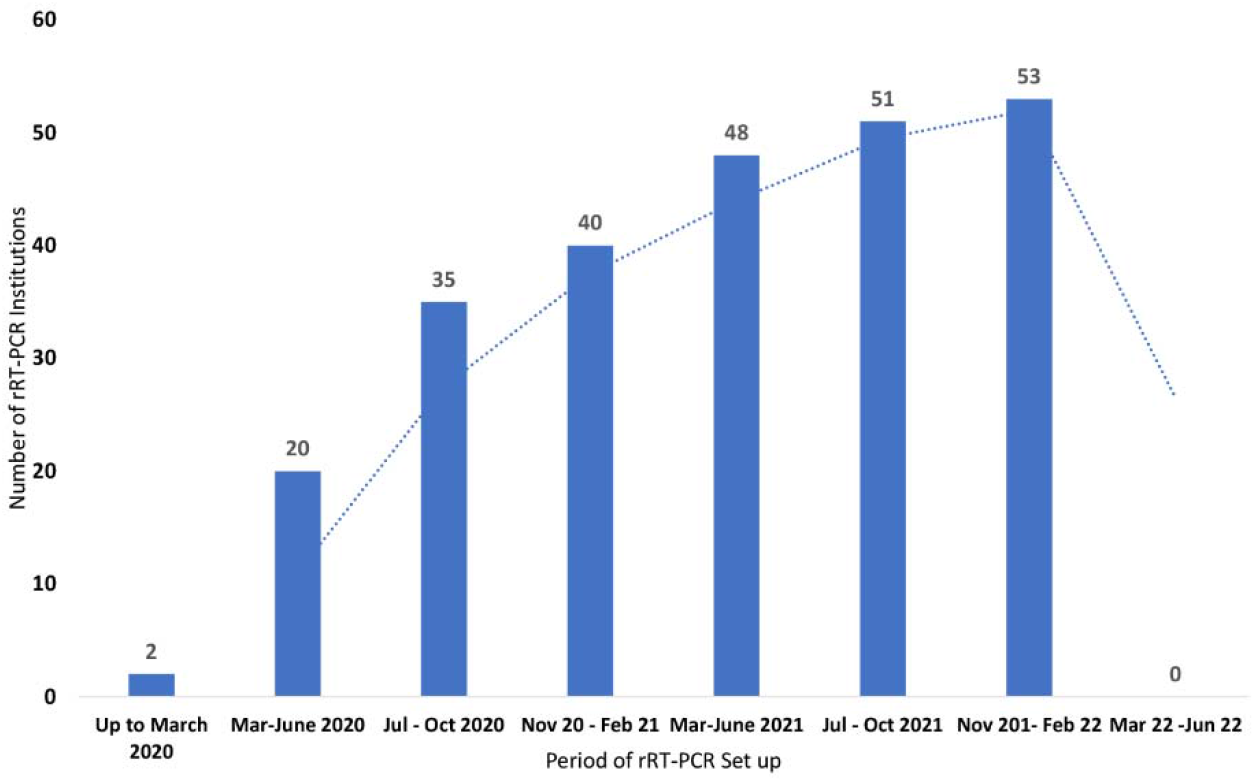
Evolution of the number of rRT-PCR testing institutions from March 2020 to June 2022.

Many (32%) of the 20 rRT-PCR equipment brands were either Sansure MA-6000 RT PCR or Bio-Rad CFX 96 DX RT PCR. Three (3) of the institutions commenced testing in mid-to-ending of March 2020. The number increased to 35 by October 2020, 40 by February, 2021; 48 by June 2021; 51 by October 2021 and 53 by June 2022. There has been no new testing institution up to July 2022. Twenty-seven, 48 (90%) of 53 testing institutions had Biosafety cabinet class II (BSCII), 3(6%) had BSC III, and 2 (4%) had glove boxes – which are essentially BSC III (***Figure 5***).

**Figure 5:**
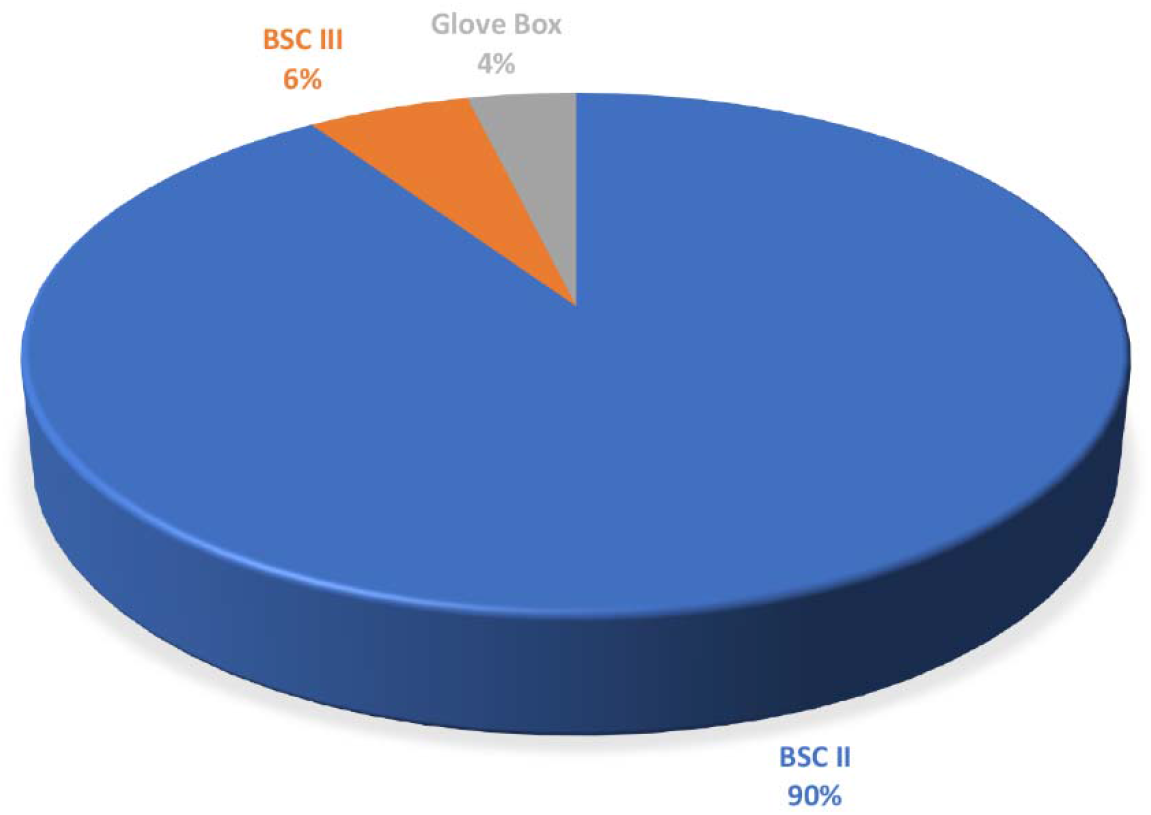
Number of each of Biosafety Cabinet (BSC) classes available in the rRT-PCR testing laboratories.

There were 25%, 96% and 100% of personal protective equipment (PPE) adequacy in 2020, 2021 and 2022 respectively. The average cost of reagents and other consumables per test, and the cost of tests to patients have decreased from GHC 180($23) (in June 2020) to GHC 112($14) (June 30, 2022) and GHC 348($45) (in June 2020) to GHC 314($40) (by June 30, 2022) respectively. Turn-around-time (TAT) for patients’ test reports decreased from an average of 26 hours in June 2020 to 16 hours in 2021 and 19 hours by June 30, 2022. The average number of tests done by each laboratory per day within the same periods were 388, 558 and 289 respectively. The institutions that had some activity or transactions with Zipline, SORMAS and PanaBios were 28%, 81% and 77% respectively. Eleven, 11(21%) of those with some transactions with Zipline received samples via them, 3 (5%) both received and dispatched samples via Zipline, and 1(2%) only dispatched samples through Zipline. Thirty-eight, 38(72%) of the institutions (all in the Greater Accra and Ashanti Regions) had no dealings with Zipline.

## Discussions

This study aimed to determine CoViD-19 rRT-PCR testing capacity in Ghana from March 2020 to July 2022, and to make some recommendations, which might help in dealing with other outbreaks such as the haemorrhagic Marburg Virus Disease (MVD) – in terms of testing. Ghana is (currently) adequately resourced for possible recurrence of CoViD-19 or Marburg virus (MV) outbreak in terms of rRT-PCR testing, however, there will be a need for redistribution of the available resources and increasing of the safety materials and equipment such as PPEs and BSCs class III across the 16 regions, to achieve success in case of such outbreak. This must be done, considering population density of each region vis-à-vis demand and supply forces. In March 2020, when Ghana recorded its first 2 cases, rRT-PCT testing was the most important requirement for the identification of infected persons, to contact tracing, isolating and quarantining, and to clinically design management strategies for infected persons.(10) Evolution of the number of testing institutions was examined in this study. Before March 2020, the Noguchi Memorial Institute for Medical Research (NMIMR), the National Public Health Reference Laboratory (NPHRL), and the Kumasi Centre for Collaborative Research (KCCR) were ‘adequately resourced’ to test for CoViD-19 virus using the rRT-PCR. By June, 2020, there were 20 CoViD-19 PCR testing institutions, mostly concentrated within the Accra metropolis of the Greater Accra Region. These had an equal private-to-public ownership ratio - with a few scattered in other major cities such as Takoradi in the Western Region and Kumasi in the Ashanti Region.

The rate with which Ghana developed this testing capacity is remarkable (***Figure 4***), and it was comparable to what had been reported in few African countries. (11) These 53 institutions exclude the majority of Ghana Health Service (GHS) and other institutions with Cepheid GeneXpert infrastructure – capable of CoViD-19 rRT-PCR testing. There are over 130 GeneXpert devices in facilities across the country,(12) often used for Mycobacterium tuberculosis testing.(13) This study found the availability of the rRT-PCR model equipment in only 9 of the 16 regions. There were none in the Ahafo, Brong Ahafo, Bono East, Central, North-East, Upper West, and Western North regions of the country. Centralizing the testing facilities to few locations negatively affects the management of any pandemic, as has been experienced in countries like Ecuador.(14) Hence, the resourcing of the GeneXperts CoViD-19 rRT-PCR specific cartridges and other biosafety requirements, will make available more than 180 facilities able to do the rRT-PCR required for CoViD-19 or MVD diagnosis. Such provisions will result in at least 3(2%) rRT-PCR facilities in the Ahafo region, 23(12%) in Ashanti, 7(4%) in Bono East, 5(3%) each in Brong Ahafo, Savannah and Western North; 10(5%) in Central, 21(11%) in Eastern, 46(25%) in Greater Accra, and 6(3%) each in Northern and Oti (***Figure 1b***). Others will include, 8(4%) in Upper East, 4(2%) in Upper West, 12(6%) in Volta, and 8(4%) in Western. Besides, there will be 17 (9%) of these testing devices available in Ghanaian research institutions, teaching hospitals, public health reference laboratories and at deprived and ‘difficult-to-reach’ locations. This will help build the capacity of local professionals through training, reduce testing cost, and reduce the TAT. It will also increase accessibility and rapid response to CoViD-19 or other viral outbreaks such as the Marburg virus - which has a higher Case fatality rate (CFR) (of 50%), compared to CoViD-19.

At the genesis of the pandemic in 2020, PPEs were scarce – even for health professionals involved in patient contact, sample collection and testing.(15) However, by June 2022, PPEs were adequate for these purposes (i.e. 25% adequacy of PPEs in 2020 compared to 100% in 2022). In case of a Marburg Virus (MV) outbreak, safe sample collection, transport and testing for these risk group 4 pathogens would require all PPEs for CoViD-19 including (K)N95 Medical masks, eye protection (goggles), gloves, disposable gowns, and face shields. In addition, there would be a need for (personal) positive pressure suits or additional protection over laboratory clothing, such as solid-front gowns with tight-fitting wrists, two pairs of latex (preferably rubber) gloves, and an approved particulate respirator (e.g., N95 or higher), and impermeable cleanable footwear with its-impermeable (disposable) covers.(16) Also, there would be the need for containment level 4 facilities with a minimum of BSC class III for most laboratory procedures, and from this study we have just a few of such BSCs, but are mostly concentrated in the Greater Accra and Ashanti regions. Thus, more of BSC class III are required. This study found a reduction in the average cost of reagents and consumables per test, and a decline in the number and cost of tests from June 2020, through to June 2022. These reductions could be as a result of increases in the number of the testing service outlets/institutions, opening of the country’s border for imports (there by increasing the supply side of resources), vaccination against the virus, reduced public restrictions, and the country being in the post-peak stage of the pandemic – all of which affect the supply and demand forces for the rRT-PCR testing. Similar findings have been reported in some African countries, including South Africa and Nigeria.(17,18)

All the participants from the public testing facilities couldn’t tell the cost of the PCR reagents and consumables per test, but most from the commercial private and public-private-partnered (PPP) institutions were able to provide such costs. The average cost of testing (reagents and other direct consumables) declined by 38% from June 2020, and 20% from June 2021. These declines in average cost of testing, and that of the test prices could be due to equipment placement, and reagent/consumable high-purchase services expanded by the players (in the industry) within the mid-2021. Other factors such as the increasing in the number of suppliers, may have contributed to these trends in prices. The average cost of tests within the same period decreased from GHC 349 ($45) in 2020/21 to GHC 314 ($40) by June 2022. It could be said from these, that an average of GHC 168 ($22) (i.e.93%), GHC198 ($25) (130%), and GHC202($26) (180%) direct-gross profits were made per test each year (ending) June 2020, June 2021 and June 2022 respectively. These findings have been collaborated by other reports from other African countries.(17,18) The average number of CoViD-19 PCR tests done by each laboratory increased from 388/day (in 2020) to 558/day (in 2021) and has declined to 289/day by June 2022. These decreases in costs and number of tests (especially from the 2^nd^ quarter of 2022) could be due to availability and increased access to vaccines, reduced infection prevalence and incidence of the virus, acquisition of immunity and likely reduced virulence of the virus (at the post-peak phase). The reduction in, and non-patronage of the Zipline services in the Greater Accra region could be due to the proximity of patients to many testing institutions within the region. It is also significant to note that, the North East region, which lacks a CoViD-19 rRT-PCR testing institution could dispatch its samples to a nearby region with these drones, since there is a drone service centre at Vobsi in the West Mamprusi Municipality of the region.

In a pandemic like CoViD-19, testing must be linked to public and community health information management systems to help in decisions on surveillance, patient management, contact tracing, and further testing. By June 30^th^ 2022, most, 43 (81%) of the testing institutions were in-putting their CoViD-19 laboratory data on the SORMAS platform to help monitor and survey some epidemiological characteristics of the CoViD-19 pandemic. This activity also helped in the management of the pandemic, as decisions were mostly based on reliable data within Ghana and by extension, West Africa. Besides, most, 41, (77%) of the institutions had PanaBios certification and validation, which gave some international permissions to travelers and states with regards to their CoViD-19 infectious status, through the PanaBios Trusted Travel platform.

This study strongly recommends that, all testing institutions dealing with pathogens of public health importance, concern, epidemic or pandemics, henceforth, must compulsorily be on surveillance platforms such as the SORMAS to help in informed surveillance and monitoring. Also, the number of locations and services of Zipline, especially in the seven regions which are currently without actual rRT PCR testing facilities need to be expanded. When this is done widely, the country could limit the number of testing institutions across the remote parts of the country – thus saving costs and reducing the risk of infections that might occur due to personal(human) delivery services and community fallouts. In addition to these, this transport technology may be employed for other health services – such as the transport of blood products for transfusion within the Ghana health system and in any infectious outbreak. Moreover, with these findings and the knowledge of CoViD-19 rRT-PCR testing techniques, resourcing the GeneXpert devices and facilities with CoViD-19 rRT-PCR reagents (or MV reagents -in MV outbreak), consumables, PPEs and other biosafety devices are recommended.

## Conclusions

Through the CoViD-19 pandemic, Ghana has built adequate capacity for molecular testing and is adequately resourced for recurrence of CoViD-19 or Marburg virus outbreak in terms of diagnosis with rRT-PCR. However, Ghana needs redistribution of these resources, satisfy additional safety requirements for Marburg virus testing and equip over 180 GeneXpert facilities with the additional infrastructure and materials required for testing. There is also a need to expand the services of infection control centers in case of hemorrhagic epidemic or pandemic. It is recommended that, all testing institutions dealing with pathogens of public health concern, henceforth, must compulsorily be on surveillance platforms such as the SORMAS to help in informed surveillance and monitoring. The CoViD-19 pandemic has brought many health, social and economic challenges. However, Ghana now has adequate testing capacity for diagnosis. There are now adequate PPEs, improved data management, technologies for transport of health resources, and an added opportunity to improve, in case of an outbreak by other viruses like the Marburg virus.

## Data Availability

All data produced in the present study are available upon reasonable request to the authors

## Abbreviations

rRT-PCR: Real-Time Reverse Transcription-Polymerase Chain Reaction
CoViD-19: SARS – COV-2 Disease 2019
MV: Marburg Virus
MVD: Marburg Virus Disease
NAAT: Nucleic acid amplification testing
PPS: Positive Pressure Suit
GHS: Ghana Health Service
WHO: World Health Organization
SORMAS: Surveillance Outbreak Response Management and Analysis System
TAT: Average turn-around-time per test per patient
CDC: Centre for Disease Control and Prevention, USA
BSC: Biosafety Cabinet
PPE: Personal Protective Equipment

## DECLARATIONS

## Acknowledgement

The authors acknowledge the efforts of the entire staff of the department of Chemical Pathology of the University of Ghana Medical School, and all the professionals who took time off their busy schedules to respond to our questionnaire, especially the reviewers of the questionnaire. The authors are also grateful to all the participating institutions, especially those who responded to phone calls on follow-ups and those who allowed them into their facilities to verify and confirm some responses.

## Authors’ contributions

ENYN was involved in conceptualization, methodology, software, supervision, validation, Writing – original draft, Writing – review & editing; JK, EKO, DD, EKA, CO & HAA were involved in methodology, supervision, software, data analysis, writing – original draft, writing – review & editing; ROA, MA, ABA, BYA, SAC & SDA contributed in validation, writing – review & editing. All authors read and approved the manuscript before submission

### Funding

This research did not receive any specific grant from funding agencies in the public, commercial, or not-for-profit sectors.

### Availability of data and materials

The datasets used and/or analyzed during the current study are available from the corresponding author on reasonable request.

### Ethical Considerations and consent to participate

Not Applicable

### Consent for publication

Not applicable.

